# Geospatial analysis reveals distinct hotspots of severe mental illness

**DOI:** 10.1101/2022.03.23.22272776

**Authors:** Janet Song, Mauricio Castaño Ramírez, Justin Okano, Susan K. Service, Juan de la Hoz, Ana M Díaz-Zuluaga, Cristian Vargas Upegui, Cristian Gallago, Alejandro Arias, Alexandra Valderrama Sánchez, Terri Teshiba, Chiara Sabatti, Ruben C. Gur, Carrie E. Bearden, Javier I. Escobar, Victor I. Reus, Carlos Lopez Jaramillo, Nelson B. Freimer, Loes M. Olde Loohuis, Sally Blower

## Abstract

**Background:** The identification of geographic variation in incidence can be an important step in the delineation of disease risk factors, but has mostly been undertaken in upper-income countries. Here, we use Electronic Health Records (EHR) from a middle-income country, Colombia, to characterize geographic variation in major mental disorders.

**Method:** We leveraged geolocated EHRs of 16,295 patients at a psychiatric hospital serving the entire state of Caldas, all of whom received a primary diagnosis of bipolar disorder, schizophrenia, or major depressive disorder at their first visit. To identify the relationship between travel time and incidence of mental illness we used a zero-inflated negative binomial regression model. We used spatial scan statistics to identify clusters of patients, stratified by diagnosis and severity: mild (outpatients) or severe (inpatients).

**Results:** We observed a significant association between incidence and travel time for outpatients (N = 11,077, relative risk (RR) = 0.80, 95% confidence interval (0.71, 0.89)), but not inpatients (N = 5,218). We found seven clusters of severe mental illness: the cluster with the most extreme overrepresentation of bipolar disorder (RR = 5.83, p < 0.001) has an average annual incidence of 8.7 inpatients per 10,000 residents, among the highest frequencies worldwide.

**Conclusions:** The hospital database reflects the geographic distribution of severe, but not mild, mental illness within Caldas. Each hotspot is a candidate location for further research to identify genetic or environmental risk factors for severe mental illness. Our analyses highlight how existing infrastructure from middle-income countries can be extraordinary resources for population studies.

## Introduction

For many diseases, delineation of geographical variations in their distribution – termed geospatial research – has played an important role in enabling the identification of specific genetic or environmental factors that contribute to their causation or course of illness (1-4). It is now widely recognized that stimulation of population-level investigations in low- and middle-income countries must be a principal priority of mental health research – both to reduce health inequities and to increase our opportunities for transformative scientific discoveries (5). With a few exceptions (6, 7), however, geospatial studies of mental disorders have been limited to upper-income countries. The work reported here extends this research field to a middle-income country, Colombia. We investigate geographic variations within the department (state) of Caldas, Colombia in the distribution of bipolar disorder (BPD), schizophrenia (SCZ), and major depressive disorder (MDD), both by individual diagnosis and by disease severity.

Caldas lies within the heart of the “Paisa” region. This region of nine million inhabitants is so-named because of the predominance there of the Paisa, one of the largest genetic isolated populations in the world (8, 9). We hypothesized that, as in some other population isolates (3), mapping of geographic variation in the frequency of mental disorders within the Paisa region might enable future research focused on elucidating the causation of these disorders. To evaluate such variation in Caldas we used electronic health records (EHRs) of the Clinica San Juan de Dios, Manizales (CSJDM), a facility that provides comprehensive psychiatric care, and for which EHRs have been recorded since 2005. CSJDM treats all patients seeking care, not limiting access by health insurance status or other socioeconomic factors, and was, until 2018, the only source of psychiatric care for Caldas, serving all of its approximately one million inhabitants.

While CSJDM was the only psychiatric facility in Caldas, we considered the possibility that geographic variation in the incidence of major mental disorders at the hospital reflect geographic accessibility to treatment, i.e. that they result from the phenomenon of distance decay whereby the proportion of individuals who seek to obtain treatment at a facility decreases with increasing distance from the facility (10, 11). To exclude this possibility, we evaluated the effect of travel time on incidence. We then conducted more detailed analyses to pinpoint hotspots (locations with an overabundance of patients), both in terms of encounters overall as well as stratified by severity and diagnosis.

## Methods

### Setting and Population

To conduct our geospatial analyses, we plotted maps of Caldas using shapefiles from the National Statistics Department of Colombia (DANE) (12). The department has 27 municipalities, with the city of Manizales, the capital municipality, comprising ∼44% of its total population.

### EHR data and Inclusion criteria

Since 2005 the CSJDM has maintained EHRs on all outpatient visits and inpatient admissions. The EHRs include both unstructured free text as well as structured data (demographic information, residential address, and diagnostic codes using the International Statistical Classification of Diseases and Related Health Problems: 10th Revision (ICD-10)(13)). The authors assert that all procedures contributing to this work comply with the ethical standards of the relevant national and institutional committees on human experimentation and with the Helsinki Declaration of 1975, as revised in 2008. All procedures involving human subjects/patients were approved by the Institutional Review Boards at CSJDM and UCLA. Specifically, approval was obtained to upload a version of the EHR database with patient names, dates (of birth and service), and ID numbers removed, to a HIPAA-compliant Amazon Web Services server and to conduct the analyses described here. Individual informed consent was not required for this study.

Using records from 2005-2018 for all adult patients 18-90 years, residing anywhere in Caldas, we focused our investigation on the 16,376 individuals assigned a diagnosis of BPD (ICD-10, F31), SCZ (ICD-10, F20), or MDD (F32 or F33) at their first visit to the hospital; we defined these as incident cases. We used these initial diagnoses in all of our analyses, in order to eliminate possible biases introduced by right censoring of diagnostic data.

To conduct geospatial analyses, we identified the subset of diagnosed individuals with available residential addresses (N = 16,308). We assigned longitude and latitude coordinates to each patient’s address (a process termed “geocoding”) using the Spanish language setting of OpenCage’s geocoder through the R package opencage (14). As input, OpenCage takes an address as free text. Prior to geocoding, we formatted addresses according to the following procedures. Most addresses (N = 11,745) fit within a grid pattern that is typical throughout Colombia, comprised of parallel streets called “calles” and perpendicular streets called “carreras”. We formatted addresses of this type in a consistent format according to the pattern “Calle [X], Carrera [Y], [Municipality], Caldas”, where X is the street number for the “calle”, and Y is the street number for the “carrera”. The remaining addresses (N = 4,563) were situated in small rural areas called “veredas” which have few roads. For these addresses we took the name of the vereda or other address entry and appended the municipality and department names as above.

The output of OpenCage is a list of potential longitude and latitude coordinates with corresponding accuracy confidence scores. If multiple potential matches were returned, the most relevant was selected, based on OpenStreetMap data (15) relevance criteria for Colombian addresses. We excluded addresses with OpenCage confidence scores lower than 6; this ensured that all addresses included in the study lie within a 7.5 km bounding box measured diagonally. This procedure yielded geospatial coordinates for 16,295 of the 16,308 patient addresses; all analyses reported here focus on these 16,295 georeferenced patient records.

### Calculating incidence of mental illness diagnoses

We calculated the annual incidence of each diagnosis, by municipality: for each year between 2005-2018 we divided the number of EHR incident cases living in the municipality by the number of inhabitants of the municipality as specified in the WorldPop database (16). For the majority of municipalities, the annual number of cases was sparse, therefore we conducted all of our geospatial analyses using the 14-year aggregate of the annual incidences.

We then used the geocoded addresses to assess incidence of the three diagnoses unconstrained by municipality boundaries. We imposed a grid (with a cell size of 5km by 5km) on the map of Caldas and counted the number of patients who resided in each cell. Incidence (at the cell level) is the number of EHR incident cases living in the cell, divided by the total number of residents in the cell. We calculated incidence per 1,000 individuals for both inpatients and outpatients across all three diagnoses, and per 100,000 individuals for each diagnosis. To visualize incidence, we generated maps using the R package ggplot2 v3.1.1 with shapefiles from the National Statistics Department of Colombia (DANE)(12).

### Incidence and travel time to the CSJDM

To determine whether geospatial variability in incidence of mental disorders within the EHR was a function of geographic accessibility to the hospital, we used the software package AccessMod5 v5.6.0 (17) to create a geographic accessibility map of Caldas. It shows the estimated travel time for a one-way trip to CSJDM from any location in the department.

We first constructed a friction surface map for Caldas, quantifying the minimum amount of time that it takes to traverse one square kilometer, using a specified mode of transportation. The map was constructed by combining geospatial data on topography (18), land cover (19), water bodies (20) and road networks (12). We then generated the geographic accessibility map by plotting the geographic location of the hospital on the friction surface map, and computing the travel time (in hours) to the hospital from the centroid of each grid cell.

We used the geographic accessibility map to determine whether incidence decreases as the travel time from CSJDM increases (i.e., whether it demonstrates distance decay (10)). To evaluate this relationship, we modelled the expected number of cases by travel time in hours, accounting for the number of individuals in each grid cell of the geographic accessibility map. We used a zero-inflated negative binomial regression from the R package pscl v1.5.5, a procedure which accounts for the large number of grid units that had zero cases as well as for over-dispersion of cases. Normal 95% confidence intervals and p-values were calculated for each relative risk (RR) based on the distribution generated from bootstrapping using the R package boot v1.3-20. We performed this computation separately for inpatients and outpatients overall and by diagnoses. We applied Bonferroni thresholds to correct for multiple testing (considering eight independent tests; for inpatients and outpatients overall, and for each diagnosis separately for inpatients and outpatients), with a significance threshold of 0.00625 (0.05/8).

We further performed two sensitivity analyses of incidence versus travel time. In one analysis, rather than using continuous travel time as a predictor, we dichotomized travel time as above or below the median, in order to evaluate potential non-linear effects of travel time on incidence. In the second analysis, we included sex and a sex by travel time interaction in the model, to evaluate possible heterogeneity in the observed relative risk by sex.

### Identification of hotspots

We searched for hotspots using the gridded map of incidence in Caldas. We then applied Kuldorff’s Spatial Scan Statistic (21) implemented in the software package SaTScan v9.6 (22). SaTScan uses a circular window to scan a gridded surface. The window varies in size from one that encompasses a single grid cell to one that contains a predefined maximum population size. Use of a varying size window enables identification of hotspots unconstrained by municipality administrative boundaries. We used a cell size of 5km by 5km and set the maximum window size to encompass 25% of Caldas’s population. The software identifies hotspots by optimizing a likelihood ratio function and estimates their statistical significance by using a Monte Carlo hypothesis procedure (21). We focused these analyses on severe mental illness only (as defined by inpatient admissions). To account for multiple testing (four different tests, all inpatients and, separately, the three diagnoses), we applied a Bonferroni correction with a significance threshold of 0.0125 (0.05/4). As a sensitivity analysis, we also performed separate hotspot analyses for males and females.

We also used SaTScan to quantify and visualize the uncertainty in defining the borders of hotspots, using Oliveira’s F function (23). This function assigns an intensity value, ranging from zero to one, to each grid cell surrounding a hotspot, reflecting the probability that the grid cell belongs to the hotspot. To visualize hotspots and the uncertainty in their borders, we used QGIS v3.10 (24). The Oliveira’s F values were colored in categories split by Jenk’s natural breaks (25, 26), a setting in QGIS that enables better visualization of potential borders. We then conducted a sensitivity analysis, exploring for each diagnosis how the identified hotspots changed in size/location when varying the maximum allowable size of the reported cluster (27).

## Results

### Study Population

The study included all individuals whose initial record in the CSJDM EHR database indicated one of three major mental illness diagnoses (SCZ, BPD, and MDD), and for whom we were able to geocode the location of their residence within a 7.5 × 7.5 km bounding box (N=16,295). For most of these addresses (71%) the assignment was even more precise (to a 0.5 km × 0.5 km bounding box). Table 1 provides a demographic overview of the study population, by diagnosis, for both inpatients and outpatients. For this dataset, as for the EHR database overall (data not shown), the ratio of BPD incidence to SCZ incidence (∼5:1) is striking, considering that, in many other countries these diagnoses display incidences that are roughly equivalent (28, 29).

**Table 1.** Demographics. Total number of patients included in study by diagnosis, severity (inpatients vs outpatients), and gender. Only subjects with complete information were included in the study. BPD=bipolar disorder, MDD=major depressive disorder, SCZ=schizophrenia

|  | Inpatients (n = 5,218) |  | Outpatients (n = 11,077) |  | All Patients (n = 16,295) |  |
| --- | --- | --- | --- | --- | --- | --- |
|  | Men | Women | Men | Women | Men | Women |
| **Total** | 2225 (43%) | 2993 (57%) | 3512 (32%) | 7565 (68%) | 5737 (35%) | 10558 (65%) |
| **BPD** | 861 (38%) | 1431 (62%) | 937 (35%) | 1756 (65%) | 1798 (36%) | 3187 (64%) |
| **MDD** | 862 (37%) | 1476 (63%) | 2157 (28%) | 5645 (72%) | 3136 (31%) | 7121 (69%) |
| **SCZ** | 863 (91%) | 86 (9%) | 418 (72%) | 164 (28%) | 803 (76%) | 250 (24%) |

### Geographic variation in the incidence of major mental disorders, by severity and diagnosis

The pattern of geographic variation in incidence, at the municipality level, differs substantially by severity of illness (as defined by inpatient vs outpatient status). For inpatient admissions (Figure 1A) the incidence does not appear to be strongly related to proximity to the hospital, and the highest incidence is in a municipality (Aranzazu), located ∼ 55 km from the hospital. This pattern reflects the high concentration of severe mood disorders in this municipality compared to the department as a whole (Figure 1B); an incidence of inpatient BPD which is nearly fourfold higher (862/100,000 vs. 219/100,000), and an incidence of inpatient MDD which is more than 1.5 times higher (364/100,000 vs. 235/100,000). For inpatient MDD this incidence is nearly as high as that in the municipality (Manizales) containing the CSJDM (387/100,000). For SCZ the incidence is highest in a cluster of three municipalities located ∼ 111 km from the hospital, where it is about twofold higher than in the department as a whole (93/100,000 vs. 45/100,000). For outpatients (Figure 2A) the pattern of variation suggests a process of distance decay; the highest incidence is in Manizales, largely due to the concentration of MDD cases in this municipality compared to the department, overall (1,171/100,000 vs. 746/100,000) (Figure 2B).

**Figure 1.**
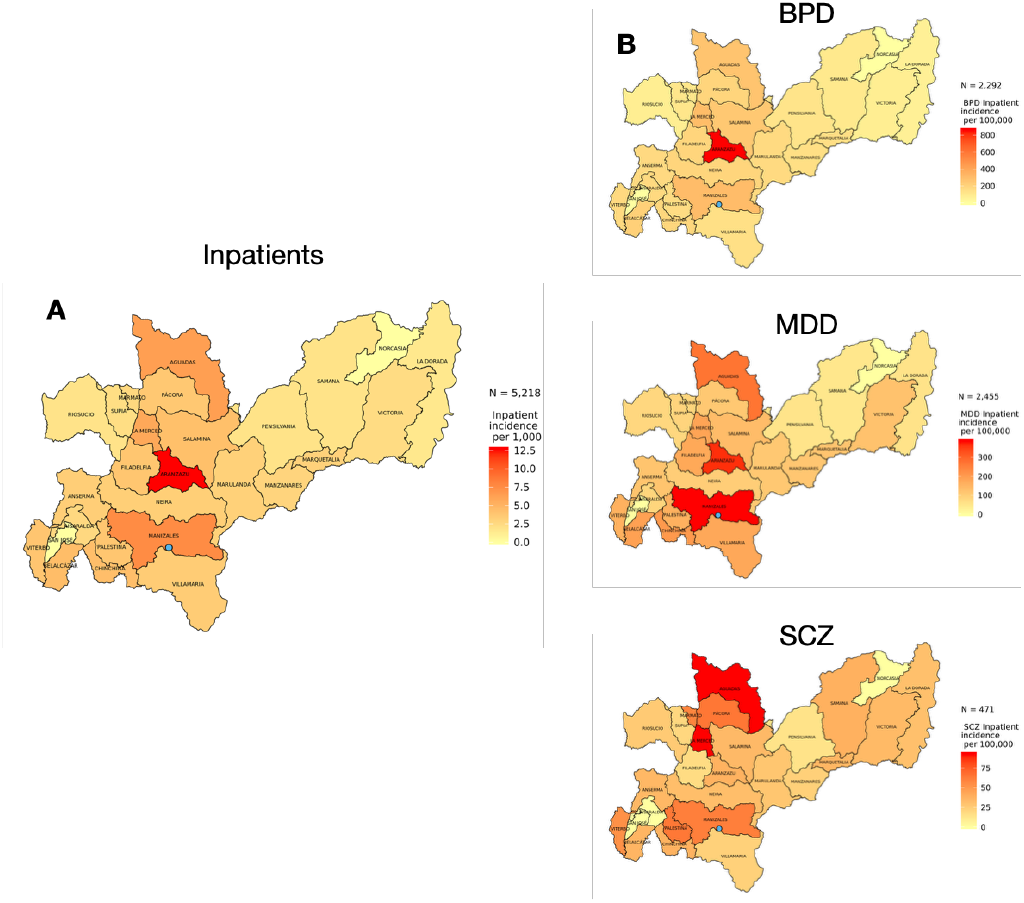
Geospatial patterns of the incidence of severe mental illness (inpatient admissions). The maps show the incidence for each municipality for patients whose first visit to the hospital was as an inpatient; (A) overall (per 1,000), and (B) stratified by diagnosis of BPD (bipolar disorder), MDD (major depressive disorder), and SCZ (schizophrenia) (per 100,000). The blue dot on the maps indicates the location of CSJDM.

**Figure 2.**
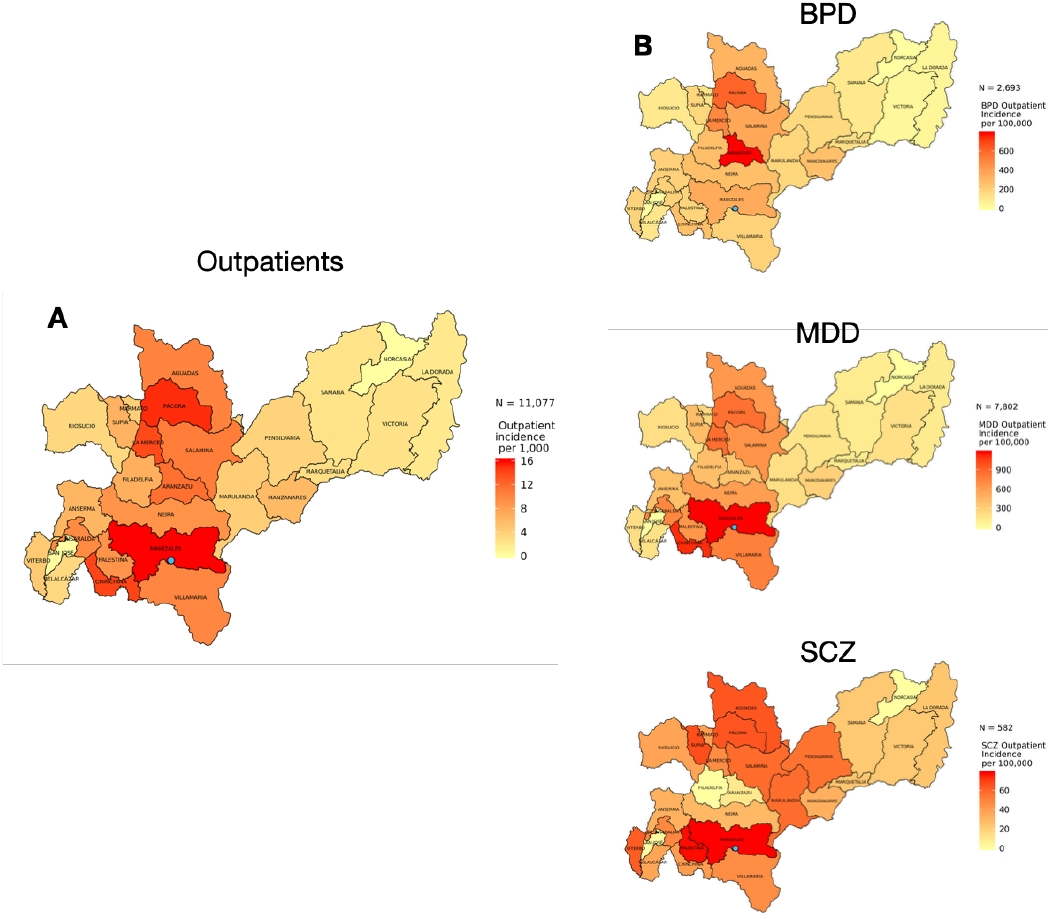
Geospatial patterns of the incidence of outpatient mental illness. The maps show the incidence for each municipality for patients whose first visit to the hospital was as an outpatient; (A) overall (per 1,000), and (B) stratified by diagnosis of BPD (bipolar disorder), MDD (major depressive disorder), and SCZ (schizophrenia) (per 100,000). The blue dot on the maps indicates the location of CSJDM.

To formally test for distance decay, we used a zero-inflated negative binomial model that included an offset for total population size in each grid (Figure S1). We modeled incidence as a function of travel time to the CSJDM using our geographic accessibility map (Figure S2), which shows that it can take more than 10 hours to reach the CSJDM from the most distant locations in Caldas. As expected from Figure 1, we found no association between the incidence of severe mental illness and travel time to the CSJDM. We did however, find a significant distance decay relationship between travel time and the incidence of outpatient visits. For every one-hour increase in travel time, the number of expected outpatient cases decreases by 20% (RR = 0.80, 95% confidence interval (0.71, 0.89), p-value = 5.67E-05, a result almost entirely driven by outpatients with MDD (Table S1). It is consistent with findings from studies conducted in upper-income countries, in which distance decay is observed mainly for mild presentations of mental illness (11, 30). We found no evidence for non-linear effects of travel time on inpatient or outpatient incidence, nor did we see any significant interactions between travel time and sex (data not shown).

### Identification of hotspots for severe mental illness in Caldas

Given the lack of significant distance decay in the distribution of severe mental illness within Caldas, we undertook analyses to delineate hotspots where these disorders show the strongest overrepresentation. We identified seven distinct hotspots (Clusters 1-7, Table 2, Figure 3), all of which show a significant overrepresentation of BPD cases, and three of which (Clusters 1, 2, and 5) cross municipality boundaries (Table 2, Figure 3A). Cluster 2 is also a hotspot for MDD while Clusters 1, 3, and 6 are hotspots for all three diagnoses (Table 2 and Figure 3 A-C). The hotspot that includes the grid cells around the hospital (Cluster 1, Table 2, Figure 3 A-C) displays the highest likelihood ratio for each of the three diagnoses. This result reflects the fact that many more patients reside in this metropolitan area than in any other region of the department; in particular, this hotspot is home to 1,476 MDD patients (92.5% of the severe MDD patients in the department) and shows the greatest overrepresentation of this diagnosis (RR = 5.47, p-value = 1.00E-17). Other hotspots, however, show patterns of overrepresentation that appear to differentiate them by diagnosis.

**Table 2.** Hotspots for severe mental illness. The rows describe the seven geographic locations (Hotspots) with statistically significant overrepresentation of cases of severe mental illness, by diagnosis. The column entitled “Cluster” is the unique cluster ID based on location and cluster size. The latitude and longitude coordinates are listed for the center of each hotspot. “N” is the number of (5km by 5km) grid units within each cluster. “LLR” is the log-likelihood ratio associated with each hotspot. “Cases Observed” is the number of cases in each hotspot, while “Cases Expected” is the number of cases expected according to a Poisson distribution in each hotspot for each diagnosis. BPD=bipolar disorder, MDD=major depressive disorder, SCZ=schizophrenia

| Cluster |  | Latitude | Longitude | N | LLR | P-value | Cases Observed | Cases Expected | RR | Population Size |
| --- | --- | --- | --- | --- | --- | --- | --- | --- | --- | --- |
| 1 | BPD | 5.041 | -75.496 | 2 | 475.87 | 1.00E-17 | 1169 | 496 | 3.78 | 227752 |
|  | MDD | 5.041 | -75.496 | 2 | 848.29 | 1.00E-17 | 1476 | 531 | 5.47 | 227752 |
|  | SCZ | 5.041 | -75.496 | 2 | 100.44 | 1.00E-17 | 242 | 102 | 3.84 | 227752 |
| 2 | BPD | 5.265 | -75.541 | 4 | 101.82 | 1.00E-17 | 111 | 20 | 5.83 | 9118 |
|  | MDD | 5.265 | -75.541 | 4 | 20.75 | 9.19E-07 | 57 | 21 | 2.72 | 9118 |
| 3 | BPD | 5.623 | -75.451 | 1 | 27.59 | 1.59E-09 | 35 | 7 | 4.90 | 3324 |
|  | MDD | 5.623 | -75.451 | 1 | 28.77 | 8.04E-10 | 37 | 8 | 4.83 | 3324 |
|  | SCZ | 5.623 | -75.451 | 1 | 16.84 | 2.11E-05 | 13 | 1 | 8.98 | 3324 |
| 4 | BPD | 5.222 | -75.798 | 1 | 15.43 | 8.61E-05 | 34 | 11 | 3.11 | 5068 |
| 5 | BPD | 5.310 | -75.136 | 1 | 12.91 | 8.25E-04 | 19 | 5 | 4.25 | 2069 |
| 6 | BPD | 5.041 | -75.631 | 1 | 12.86 | 8.57E-04 | 25 | 7 | 3.40 | 3409 |
|  | MDD | 5.041 | -75.631 | 1 | 10.53 | 7.22E-03 | 24 | 8 | 3.04 | 3409 |
|  | SCZ | 5.041 | -75.631 | 1 | 10.42 | 6.37E-03 | 10 | 2 | 6.69 | 3409 |
| 7 | BPD | 5.399 | -75.181 | 1 | 11.14 | 4.03E-03 | 31 | 12 | 2.69 | 5337 |

**Figure 3.**
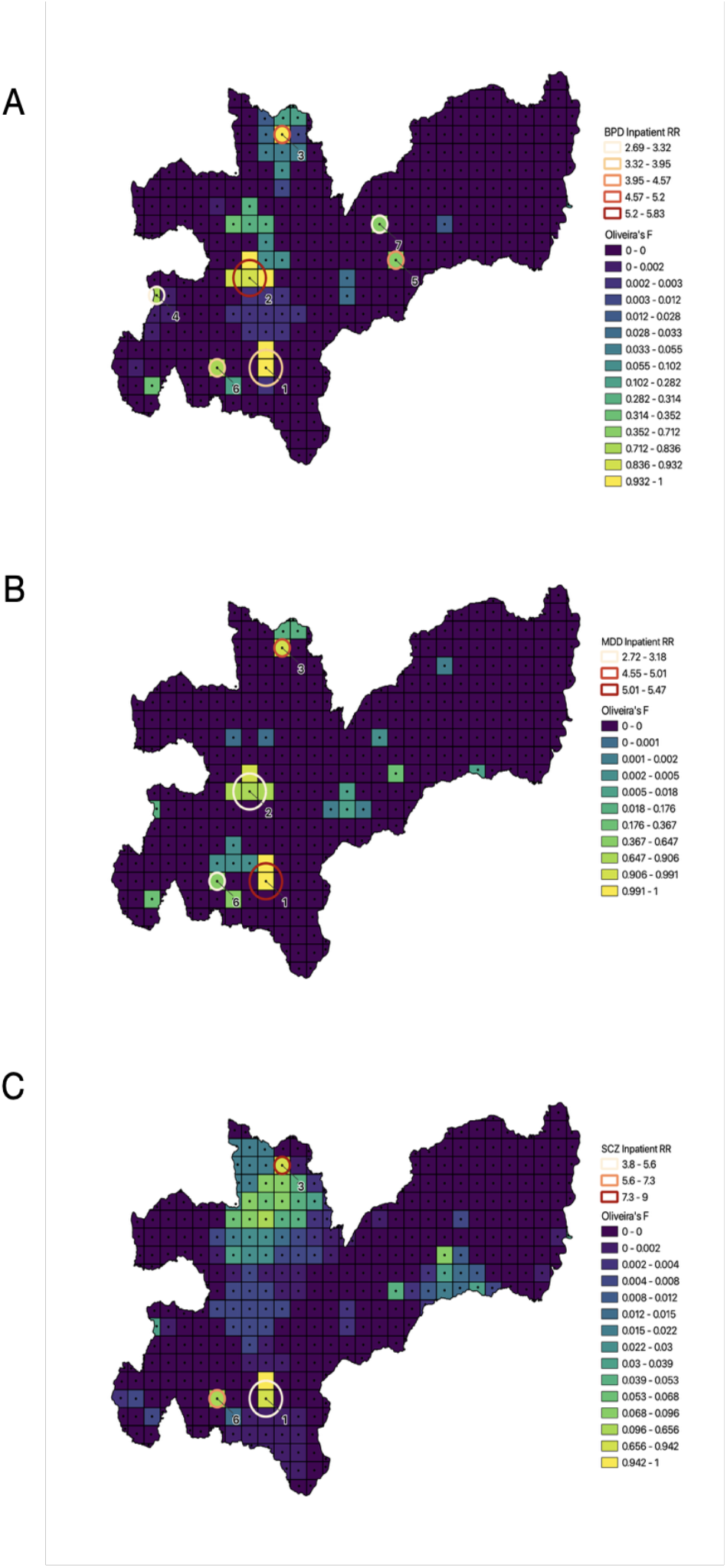
Hotspots of severe mental illness. Hotspots are shown for inpatient BPD (bipolar disorder, A), MDD (major depressive disorder, B) and SCZ (schizophrenia, C). The statistically significant locations are indicated by circles: circle size indicates cluster size and color codes correspond to different values of relative risk. The grid units are colored by Oliveira’s F values.

Cluster 2 denotes the hotspot comprising the most extreme overrepresentation of BPD cases (RR = 5.83, p-value = 1.00E-17, Table 2 and Figure 3A), is the largest of the BPD hotspots in terms of geographical extent (covering four grid units), crosses the administrative boundaries of two municipalities (Aranzazu, the municipality with the highest overall and BPD incidence [Figure 1B] and Filadelfia), and is located about 1.5 hours driving time from the hospital. The average annual incidence of severe BP at this hotspot (8.7/10,000) is among the highest frequencies worldwide (28, 29, 31),. This location may constitute a hotspot specific for severe mood disorders, as indicated by the overrepresentation there of MDD but not SCZ cases. Cluster 3, by contrast, appears to be a hotspot for severe mental illness overall, as evidenced by the overrepresentation of all three diagnoses, despite its location, four hours distant from the hospital. While the total number of inpatient cases from this location is relatively small (N=85), the nearly nine-fold overrepresentation of SCZ cases is particularly striking (RR = 8.98, p-value = 2.11E-05, Table 2 and Figure 3C). The hotspots for inpatients overall are displayed in Figure S3.

The border analysis allows us to explore uncertainty at the borders of hotspots using Oliveira’s F, however, the cluster detection algorithm is sensitive to the maximum scanning window size parameter. We explored how the identified hotspots changed in size/location when we varied the maximum allowable size of the reported cluster (Figure S4). For MDD and SCZ we observe nearly identical hotspots, irrespective of the parameters used. For BPD, when we increased the maximum reported cluster size above 40% of the population, Cluster 2 disappears. This is a consequence of the process by which SaTScan pre-computes the most likely hotspot centered at each grid, and then iteratively selects non-overlapping hotspots with the highest likelihood ratio; at settings above 40% the hotspot centered at Cluster 2 includes the area of Cluster 1. Since Cluster 1 has a higher likelihood ratio, Cluster 2 cannot be selected. The overall consistency of the hotspot patterns in these sensitivity analyses indicates the robustness of our findings. Our findings are also consistent when analyzed separately by sex; no new hotspots emerged and we identify the same clusters described in Table 2.

## Discussion

In this study we have shown the utility of a longitudinal EHR database from a middle-income country for geospatial investigations of mental illness. By using geolocated addresses to analyze diagnostic data from 16,295 patients contained in the EHR database of the single psychiatric hospital that serves the entire department of Caldas, Colombia, we characterized the geographic variation of BPD, SCZ, and MDD incidence across the department. Using geostatistical methods, we find that distance decay is statistically significant for mild but not severe presentations of mental illness, and severe mental illness concentrates in seven distinct hotspots, one of which has among the highest frequencies of BPD worldwide.

Investigations of geographic variation in the distribution of mental disorders have relied on two main approaches. Studies conducted in treatment facilities access detailed clinical information, but factors affecting access to treatment, including distance decay (10, 11, 30, 32) may cause patient samples to be unrepresentative of the population. In contrast, epidemiologic surveys provide a representative sample of the population, but usually collect only sparse, cross sectional clinical information, and depend on self-report measures which are subject to biases in recall (33) and response (34).

The CSJDM EHR database overcomes disadvantages of both approaches. The clinical data that it incorporates are extensive and longitudinal. The lack of significant distance decay in the CSJDM EHR for inpatient encounters supports our assumption that the database is representative of severe mental illness across the entire department of Caldas, and motivates the use of this EHR for population-level investigations of such illness. However, the fact that the EHR displays distance decay for outpatient encounters indicates that geographic accessibility is a factor driving utilization of the hospital for mild mental illness.

The classification of severe mental illness has traditionally subdivided it according to the three diagnoses evaluated in our study (35). Mounting evidence has called into question the validity of this classification system by documenting extensive phenotypic and genotypic correlations across these diagnostic categories (36, 37). Other lines of evidence, however, including diagnosis-specific clinical features and genetic associations, suggest that these classifications remain useful (38). These prior lines of evidence reinforce the utility of comprehensively cross-diagnostic investigations. To our knowledge, this is the first report of high-resolution geospatial analyses based on unified, unbiased ascertainment of cases for all three of these major categories of severe mental illness (39-41). Our results support both overlapping and distinct signatures across severe mental illness; while three of the seven hotspots that we identified show significant overrepresentation of cases across all three diagnoses, the most extreme hotspot (denoted by Cluster 2) displays significant overrepresentation of BPD (and to a lesser extent MDD), but not SCZ.

The hotspots that we identified may represent the most extreme manifestation of genetic or environmental risk factors for severe mental illness that are highly geographically variable in their distribution, and that might explain the broader patterns seen in the EHR database. For example, at Cluster 2 we found an incidence of BPD that is among the highest ever reported. This result should be considered in the context of the approximately five-fold overrepresentation in the database of BPD relative to SCZ, consistent with our observations in a prior study, which were based on diagnostic interviews among research participants recruited from the database (42). To our knowledge an imbalance of this extent has not been reported previously; studies in other countries have consistently described similar frequencies of these diagnoses among hospitalized individuals (28, 29).

The results reported here present a starting point for future research aimed at identifying specific genetic or environmental risk factors that contribute to geographical variation in the incidence of mental disorders within the Paisa region. The Paisa population offers unusual opportunities for such research. In the demographic history of some large genetic isolates, a process of migrations followed by rapid population expansion has created a series of geographically localized sub-isolates, in which even deleterious genetic variants can attain a sizable frequency; this history facilitates the discovery of genetic variants that have a high impact on disease phenotypes. The localized enrichment in Finland of the mutations responsible for a wide range of diseases (3) is the most well-known example of this process. The population of Caldas represents a similar kind of sub-isolate within the overall Paisa region (43).

Exposure to violent conflict, and forced displacement associated with such exposure, are environmental factors that are known to contribute to geospatial variability in disease risks. Colombia’s internal armed conflict led to the forced displacement of nearly 6 million people between the 1960s and the 2000s, one of the highest rates of any country, worldwide (44, 45). Previous studies in Colombia have linked exposure to violent conflict to poor health outcomes generally (45), and increased rates of mental health disorders, specifically (44, 46). In our data, in contrast to the hotspot at Cluster 2 discussed above, the hotspot that includes CSJDM (Cluster 1) displays a roughly similar overrepresentation of all three of the severe mental illness diagnoses. This hotspot covers a sizable city which experienced considerable migration from less urbanized areas of Caldas, and other parts of the Paisa region during the period of internal armed conflict. We speculate that further geospatial mapping studies in Caldas that incorporate information on prior migration and exposures to violence could provide unique insights into the impact of such variations in environmental exposures on the distribution of mental disorders. The analyses needed to perform these investigations are not feasible using the current dataset, which does not include the location of individuals’ residence prior to their first recorded hospital visit.

An additional limitation of our study is that, by focusing only on initial presentation to the hospital, the analyses we have performed do not account for variability in disease trajectories. Specifically, many individuals experience a change in severity (their initial visit is as an outpatient but they subsequently have at least one inpatient admission) or in diagnosis (for example, they receive an initial diagnosis of MDD and later convert to a diagnosis of BPD). Future analyses that incorporate such trajectory information may reveal different geospatial patterns than those reported here.

With a few notable exceptions (6, 7), previous geospatial analyses of mental disorders have been conducted in upper-income countries. The work reported here extends this research field to a middle-income country that has recently experienced extensive and traumatic social disruptions. Despite such disruptions, Colombia has established infrastructure, such as the CSJDM EHR database, that constitute extraordinary resources for population mental health studies. We believe that global efforts to understand the causes and trajectories of severe mental illness will increasingly rely on data resources in low and middle-income countries, and the efforts of investigators in these countries who have expert knowledge of these resources.

## Data Availability

NA

## Acknowledgements

This work was supported by R01MH113078 (to CEB, CLJ and NBF), R01MH123157 (to LMOL, CLJ and NBF), R00MH116115 (to LMOL) and R56 AI152759 (to SB). We thank Anjene Musick for valuable feedback on the manuscript.

