## Supplement for "Geospatial analysis reveals distinct hotspots of severe mental illness"

**Supplemental items included:**

Table S1: The effect of travel time on the incidence of severe and mild mental illness
Figure S1: Administrative features and population density of Caldas
Figure S2: Geographic accessibility map of Caldas
Figure S3: Hotspot and Oliveira F values for inpatients overall
Figure S4: Results of sensitivity analyses for hotspot identification

| Encounter | RR | 95% confidence interval | p-value |
|---|---|---|---|
| Inpatient | **0.88** | 0.80 - 0.97 | 1.25E-02 |
| Outpatient | **0.80** | 0.71 - 0.89 | 5.67E-05 * |
| BPD Inpatient | **0.88** | 0.79 - 0.99 | 3.61E-02 |
| BPD Outpatient | **0.84** | 0.74 - 0.96 | 1.17E-02 |
| MDD Inpatient | **0.93** | 0.84 - 1.03 | 1.76E-01 |
| MDD Outpatient | **0.79** | 0.70 - 0.88 | 5.43E-05 * |
| SCZ Inpatient | **1.05** | 0.92 - 1.19 | 5.02E-01 |
| SCZ Outpatient | **1.00** | 0.88 - 1.12 | 9.55E-01 |

**Table S1. The effect of travel time on the incidence of severe and mild mental illness**

Relative Risk (RR), 95% confidence interval, and p-values for a zero-inflated negative binomial model. We find the effect for each hour increase in travel time on the expected incidence overall and for individual diagnoses, in both cases considering inpatients (severe illness) and outpatients (mild illness) separately. P-values with an asterisk were significant after Bonferroni correction for analysis of eight tests P<0.0062 (0.05/8). BPD=bipolar disorder, MDD=major depressive disorder, SCZ=schizophrenia

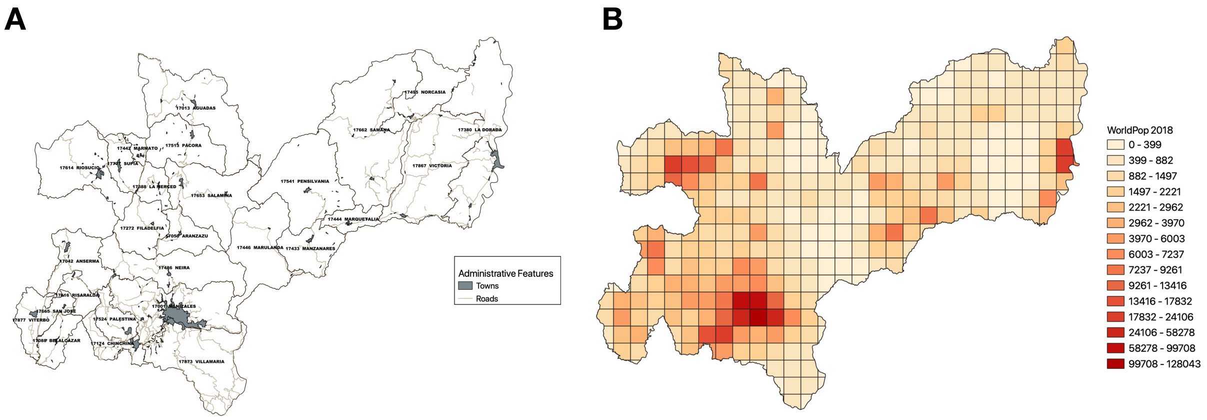


**Figure S1. Administrative features and population density of Caldas.**
The maps show main roads and towns in Caldas[1] (A) and population size estimates for each 5x5km grid as specified in the WorldPop database[2](B).

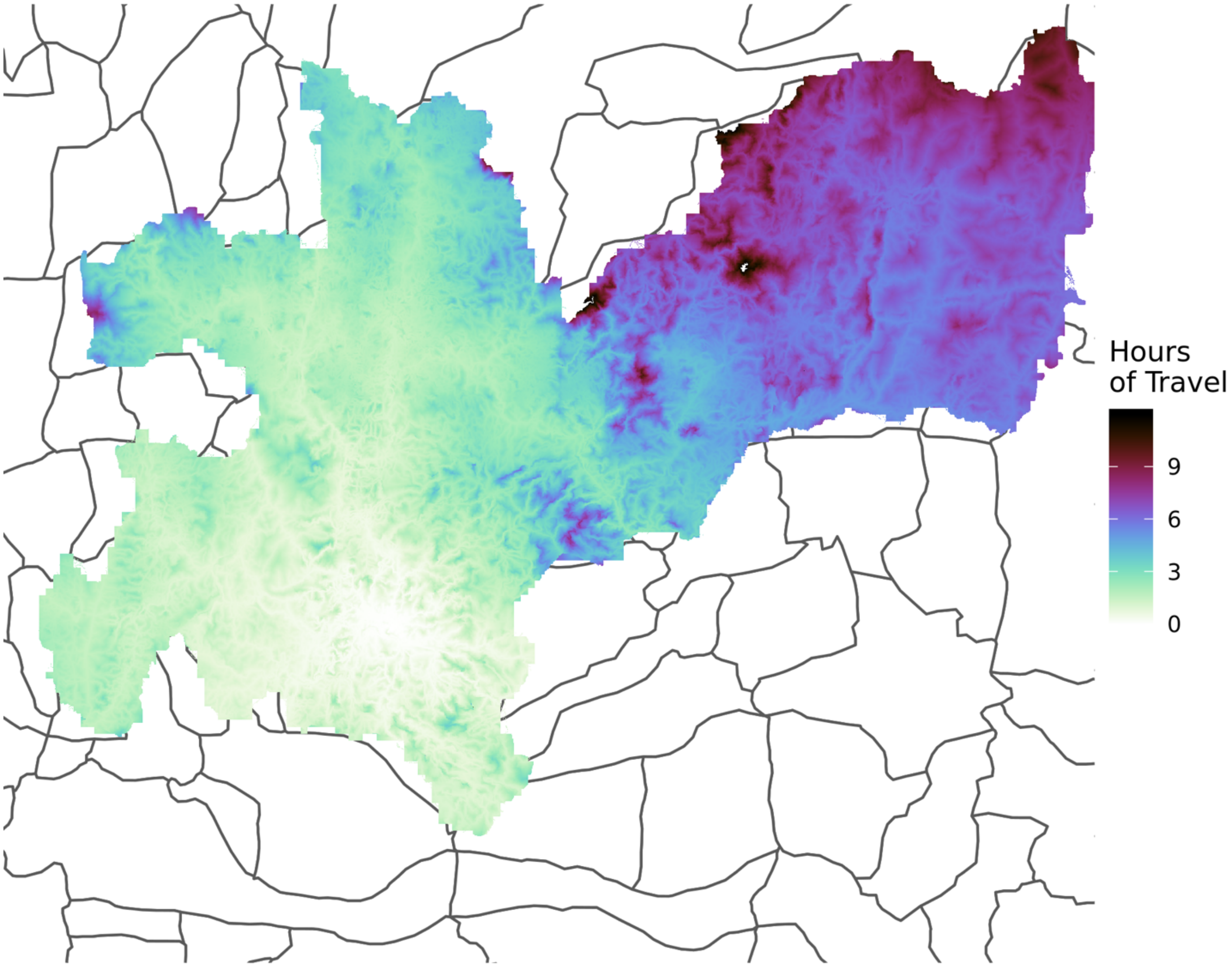


**Figure S2. Geographic accessibility map of Caldas.**
This map shows travel time to CSJDM (in hours) from anywhere in the department.

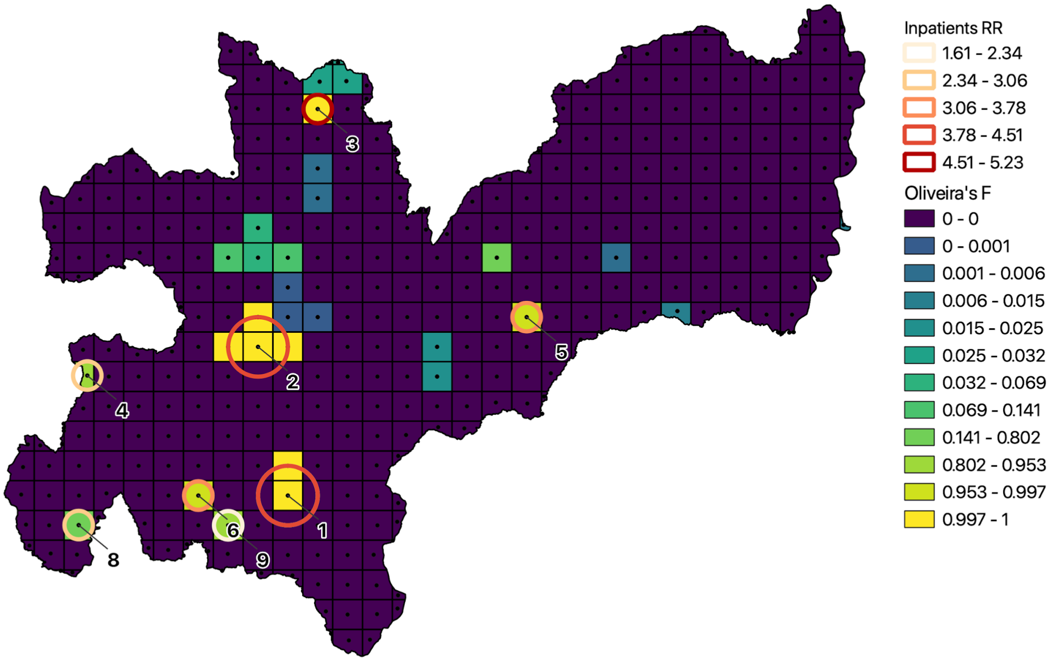


**Figure S3. Hotspot and Oliveira F values for inpatients overall.**
The statistically significant locations are indicated by circles: circle size indicates cluster size and color codes correspond to different values of relative risk. The grid units are colored by Oliveira’s F values.
